# Management of very severe tungiasis cases through repeated community-based treatment with a dimeticone oil formula: a longitudinal study in a hyperendemic region in Uganda

**DOI:** 10.1101/2023.09.15.23295530

**Authors:** Hannah McNeilly, Francis Mutebi, Marlene Thielecke, Felix Reichert, Mike B. Banalyaki, Rebecca Arono, George Mukone, Hermann Feldmeier

## Abstract

Tungiasis (sand flea disease) is a neglected tropical disease that is endemic in sub-Saharan Africa and Latin America. Tungiasis causes pain, mobility restrictions, stigmatisation, and reduced life quality. Very severe cases with hundreds of sand fleas have been described, but treatment of such cases has never been studied systematically.

During a larger community-based tungiasis control programme in a hyperendemic region in Karamoja, northeastern Uganda, 96 very severe tungiasis cases were identified and treated with the dimeticone formula NYDA®. They were repeatedly followed-up and treated again when necessary. The present study traces tungiasis frequency, intensity, and morbidity among these 96 individuals over two years.

At baseline, very severe tungiasis occurred in all age groups, including young children. Throughout the intervention, tungiasis frequency decreased from 100% to 25.8% in this group. The overall number of embedded sand fleas in this group dropped from 15,648 to 158, and the median number of embedded sand fleas among the tungiasis cases decreased from 141 to 4. Walking difficulties were reported in 96.9% at the beginning and in 4.5% at the end of the intervention.

Repeated treatment with the dimeticone formula over two years was a successful strategy to manage very severe cases in a hyperendemic community. Treatment of very severe cases is essential to control the spread and burden of tungiasis in endemic communities.

## Introduction

Tungiasis is a neglected zoonotic skin disease caused by the female sand flea *Tunga penetrans*, which burrows into the skin of the feet and other body parts that come into contact with infested soil [1-3]. Tungiasis causes significant pain, itching, and mobility restrictions, and can lead to disability and mutilation of the affected body parts [3,4]. It is endemic in impoverished populations of Sub-Saharan Africa and Latin America [5-7]. Like other neglected tropical skin diseases, tungiasis is a highly stigmatized condition [8,9]. As endemic communities lack access to effective and safe treatment, manual removal of sand fleas with non-sterile sharp objects like pins or thorns is the most common treatment method [10,11]. This practice leads to painful wounds and risk of bacterial and viral infections [12,13].

The intensity of tungiasis has previously been classified as mild (1-5 embedded sand fleas), moderate (6-30), and severe (>30) [14]. In areas with a high prevalence of tungiasis, however, very severe cases with hundreds of embedded sand fleas have been described [14-19] that are not reflected in this classification. To selectively investigate such cases, we have introduced ‘very severe’ tungiasis as an additional category, defined as presence of ≥100 embedded sand fleas (including vital, dead, and manipulated parasites). Previous studies have shown that intensity of tungiasis correlates with morbidity [4,16]. Potentially life-threatening consequences of very severe tungiasis are abscesses, septicaemia, severe malnutrition, cachexia, and anaemia [15,18,20]. As severe tungiasis causes physical disability and social stigmatisation [8,21,22], highly affected individuals are at high risk of social isolation and inability to work, which traps them in a poverty cycle.

The topical application of a formula of two dimeticone oils with different viscosities and high creeping properties (NYDA®) has been shown to be a highly effective and safe treatment for tungiasis, as it kills embedded sand fleas by clogging their respiratory, intestinal, and reproductive systems [23-25]. We describe the regression of very severe tungiasis among 96 patients who were treated with the dimeticone formula in Karamoja, northeastern Uganda [19].

## Methods

### Study site and Study population

This study took place in three parishes of Ngoleriet sub-county, Napak district, Karamoja sub-region, northeastern Uganda [19]. In the study area, people dwelled in simple huts made of sticks with earthen floors and mainly existed on pastoralism and small-scale crop farming. Hunger was common and most people were illiterate [19]. At baseline, the overall prevalence of tungiasis was 62.8% among 4,035 examined residents in the study area [19]. The present study presents a spotlight on 96 very severe tungiasis cases at baseline and follows this group over two years, during which re-occurring tungiasis lesions were systematically detected and treated.

### Case detection and Intervention

The 96 cases reported here were identified during the implementation of a larger community-based One Health tungiasis intervention, based on systematic rounds of topical treatment of tungiasis cases with the dimeticone formula NYDA®, accompanied by community information about tungiasis treatment and prevention [19]. Between February 2021 and December 2022, eight systematic rounds of tungiasis case detection and treatment were conducted among the entire population of the study area (approximately every three months). A team of 24 Village Tungiasis Health Workers (VTHWs) washed the community members’ feet with soap and water, and visually examined the feet and other affected body parts. Subsequently, they treated tungiasis cases with the dimeticone formula. The VTHWs were bilingual in the local language NgaKarimojong and English and had been specifically trained for this task. During their house-to-house visits, the field team further promoted preventive measures, such as keeping the feet and house floors clean.

The very severe cases were treated by applying the dimeticone formula NYDA^®^ to the whole foot (and other affected areas in ectopic cases) and rubbing it into the skin. Treatment was repeated every other day until no live sand fleas could be detected. Patients were encouraged to contact the VTHW whenever they noticed new sand flea lesions, as to receive treatment with dimeticone formula in between the systematic treatment rounds. The numbers of applications per treatment and of inter-round treatments among the 96 individuals were not recorded.

### Data collection and Data analysis

Clinical examinations were carried out in a door-to-door approach by the team of VTHWs as part of the community-wide intervention. They also recorded demographic data and asked people with tungiasis to categorise pain and itching (none/only a little/quite a lot/very much) and if they had difficulty walking (yes/no). VTHWs used mobile phones to record their answers in English in the Open Data Kit (ODK Inc., San Diego, CA, USA).

Data were transferred to Microsoft Excel files (Excel® for Microsoft 365 MSO, Version 2307) and were double-checked by the project’s data manager. Data analysis was carried out using IBM SPSS Statistics (Version 27). Descriptive statistics include median, percentages, ranges and IQR.

### Ethics

The VTHWs informed all participants in their local language about the aims and methods of the study, answered any questions, and collected written consent, either in the form of a signature or fingerprint. In the case of children, the caretaker was asked for written consent and the minor for verbal assent. Ethics approval for this study was given by the Vector Control Division of the Ugandan Ministry of Health Ethical Committee (VCDREC112/UG-REC-018) and the Uganda National Council of Science and Technology (HS2623, July 2019).

## Results

At baseline, 53 of the 96 very severe cases (55.3%) were children, including 22 young children ≤ 5 years (22.9%); and 24 were elderly ≥60 years (25.0%). The median age was 12 years (range: 6 months-87 years, IQR 6-59 years). Most of the very severe cases were female (60.4%, n=58), mirroring the high proportion of females (63.5%) among the overall study population [19]. Most of the very severely affected children at baseline were boys (60.4%, n=32). The very severely affected adults were predominantly women (86.0%, n=37).

Although the 96 very severe cases only represented 2.4% of the examined overall population (n=4,035), at baseline they carried 26.6% (15,648/58,806) of all embedded sand fleas in the community (Table 1). The lesions among the 96 individuals were predominantly located in the feet. However, in 61 cases (63.5%), tungiasis was also found in ectopic sites (i.e., other than the feet), mainly on the fingers and palms. In few cases, the elbows (n=3) and knees (n=3) were also affected. All 61 individuals with ectopic tungiasis also had tungiasis lesions in their feet. Clinical examination further showed that all 96 individuals presented signs of local inflammation (i.e., redness, warmth, oedema, and pain). Pus or abscesses were present in 51.0% (n=49). At baseline, the morbidity was very high. Almost all the 96 very severe cases had difficulty walking (96,9%), 80.2% reported that they experienced ‘very much’ pain, and 77.1% said they suffered from ‘very much’ itching.

**Table 1:** Regression of tungiasis among the 96 individuals with very severe tungiasis infection at baseline.

| Year | Treatment round | Tungiasis present (N=96) | Proportion of very severe cases (N=96) | Median number of embedded sand fleas* among cases (range; IQR) | Total no. of embedded sand fleas* |
| --- | --- | --- | --- | --- | --- |
| 2021 | 1 (Baseline) | 100% (96/96) | 100% (96/96) | 141 (101-591; 115-186.5) | 15,648 |
|  | 2 | 87.5% (63/72) | 4.2% (3/72) | 13 (2-282; 6-29) | 1,717 |
|  | 3 | 65.4% (51/78) | 9.0% (7/78) | 24 (1-220; 9-50) | 2,056 |
|  | 4 | 63.6% (42/66) | 4.5% (3/66) | 15 (1-167; 5-41) | 1,226 |
| 2022 | 5 | 47.9% (23/48) | 0% (0/48) | 12 (1-79; 7-19) | 386 |
|  | 6 | 23.9% (16/67) | 0% (0/67) | 14.5 (1-86; 1-31) | 357 |
|  | 7 | 39.3% (22/56) | 1.8% (1/56) | 3.5 (1-103; 2-11) | 260 |
|  | 8 | 25.8% (17/66) | 0% (0/66) | 4 (1-31; 1.5-17) | 158 |

\*includes vital, dead, and manipulated sand fleas

In the subsequent treatment rounds, between 78 (81.3%) and 48 (50.0%) of the 96 individuals could be followed up (Table 1). The notes from the VTHWs show that many of the absences were due to people having travelled to other villages for casual work or visiting family, or that they were herding cattle elsewhere.

Throughout the intervention, the frequency of tungiasis, the median number of lesions among tungiasis cases, and the total number of lesions decreased considerably among the 96 individuals (Table 1). While re-occurrence of tungiasis was common, particularly during the first year of the intervention, the intensity and overall burden of disease decreased considerably. Table 1 shows that at baseline the median number of embedded sand fleas was 141, and it decreased to 12-24 in rounds 2 to 6, which reflects moderate intensity [14]. In rounds 7 and 8, the median had dropped to 4, which corresponds with mild intensity [14]. Re-occurrence of very severe tungiasis among the 96 individuals was rare in year 1 (between 3 and 7 cases per treatment round), and in all of year 2, only one relapse of very severe tungiasis was found in this group (Table 1). This relapse case was a woman in her fourties with 103 sand flea lesions in her feet, who had had re-occurring tungiasis of varying intensities in all follow-up rounds, except for rounds 2 (no tungiasis) and 4 (absent).

At the end of the study (treatment round 8), the frequency of tungiasis cases among the 66 examined individuals had dropped to 25.8% (n=17) (Table 1). Between them they had a total of 158 embedded sand fleas, which represents a reduction of 99% compared to 15,648 at baseline. In round 8, the frequency of tungiasis among the 35 examined children had decreased to 17.1% (5 boys and 1 girl), while among the 17 adults ≥ 60 years, 41.2% had tungiasis (6 women and 1 man; data not shown in Table 1). At this time, ‘very much’ pain was reported by 6% (n=4), ‘very much’ itching by 4.5% (n=3), and walking difficulties by 4.5% (n=3).

### Case studies

One of the very severely affected patients was an infant with 114 tungiasis lesions on the feet, 39 of which had been manipulated with sharp objects. Embedded sand fleas were located on all toes, and covered both soles, heels, and sides of the feet. The parent and the infant’s three siblings all had tungiasis, and the eldest sibling was also very severely affected. Over the seven systematic follow-up rounds, the child remained free of tungiasis, except for round 6, when the VTHW found and treated three embedded sand fleas on the toes of her left foot, none of which had been manipulated with a sharp object. In year 2 of the study, she was walking and running normally.

The most severe case in the cohort was a woman in her fourties who lived alone. She had 591 tungiasis lesions on her body, 471 of which were located on the feet, with all toes, both soles and heels, and the lateral rims of the feet being affected. She further had 120 tungiasis lesions in ectopic sites: on all fingers (66 lesions), both palms (41 lesions), and both knees (13 lesions). At baseline, 211 sand flea lesions had been manipulated with sharp objects, and the woman reported ‘very much pain’, ‘very much itching’ and difficulty walking. In all subsequent systematic follow-up rounds, this woman was again found to have embedded sand fleas. However, the number of lesions was reduced, and ectopic sites were affected only in treatment rounds 3 and 4. In treatment rounds 6 and 7, she only had 4 and 9 embedded sand fleas, respectively, with no signs of manipulation. During the last treatment round, she was absent and the VTHW noted “went to [neighbouring sub-county] to visit relatives”.

## Discussion

Our study shows that community-wide systematic case detection and treatment with the dimeticone formula NYDA® was a successful strategy to manage very severe tungiasis cases in a setting where tungiasis was hyperendemic and living conditions were desolate. The finding that re-infections declined over time corroborates the assumption that the systematic occlusion of the genital tract of embedded sand fleas prevents eggs from being expelled to the ground and, by consequence, has a deleterious effect on the off-host life cycle of *T. penetrans*.

In our study, very severe cases of tungiasis occurred across all age groups, even among very young children. The occurrence of very severe tungiasis in children has previously been reported in case studies from Tanzania [18,20] and Brazil [17]. A study on tungiasis in children in Kenya showed that reported life quality reduction included sleep disturbances, difficulty to concentrate, feelings of shame, difficulty to walk and play, and social exclusion [4]. As disease intensity correlates with the impact on life quality [4], we assume that very severe cases of tungiasis experience the most severe life quality reduction. At young age, the tungiasis-associated walking difficulties, strong pain and itching, as well as social exclusion and co-morbidities like anaemia and malnutrition, can delay or seriously impede normal child development [26,27]. Our case presentation of the infant highlights that manual extraction of sand fleas with sharp instruments was even carried out in very small children, with risk of psychological trauma and viral and bacterial infection [12,13].

Our finding that a small minority (2.4%) of the population carried a disproportionally large share (26.6%) of the total number of embedded sand fleas is corroborated by a study from a Brazilian fishing village, where 8% (n=23) of the overall study population (n=620) carried 54.8% of the sand fleas [14]. As sand fleas expel eggs to the ground whilst embedded in the skin [3] and non-embedded adult fleas can move from one person to the next [28], individuals with very severe tungiasis must be considered ‘superspreaders’ in their surroundings.

Treatment with the dimeticone formula NYDA^®^ has previously been shown to be a safe, easy-to-use, and effective treatment for tungiasis in a proof-of-concept study in Kenya [24]. When single drops are applied to embedded sand fleas, they die off in almost 100% [25]. Our study demonstrates that the dimeticone formula is also effective in very severe tungiasis cases. However, re-occurrence of tungiasis was common, especially during the first year of the intervention, although new cases were considerably milder. A study from Madagascar showed that the application of a plant-based insect repellent (Zanzarin®) in very severe tungiasis cases reduced the re-infection rate by 89% compared to 41% when using closed shoes [16]. Thus, the application of insect repellent after topical treatment with the dimeticone formula could help to prevent re-occurrence of tungiasis in a hyperendemic area.

Elsewhere, our study team highlighted that domestic animals did not play a major role for tungiasis transmission in our study area, as few animals were present in the villages and tungiasis prevalence in animals was much lower than in humans [19]. Nevertheless, animals (namely pigs, cats, and dogs) have been shown to be important reservoirs for *T. penetrans* in other regions [29-31], and thus a One Health approach is generally recommended for tungiasis control in endemic communities.

A limitation of this study is that the number of inter-round treatments of the 96 individuals was not recorded. This is because the focus on management of very severe cases only emerged after the start of the project. Another limitation is the loss of participants after baseline due to absence from the villages. This can be explained by the fact that many had temporarily left for casual labour and to visit family, and that boys and men had gone to herd cattle. Their absence from the villages can thus be understood as a sign of increased mobility resulting from successful tungiasis treatment.

In conclusion, the key messages of our study are:

- Very severe tungiasis cases occur across the different age groups, including among young children.
- Repeated community-based treatment with dimeticone formula NYDA^®^ effectively reduces tungiasis frequency, intensity, and morbidity even in very severe cases.
- Treatment of very severe cases is essential to control the spread and burden of tungiasis in endemic communities.
- As re-infections after treatment of very severe cases are common, tungiasis control programmes should consider additional tungiasis prevention measures and adopt a long-term approach.

## Data Availability

All data produced in the present study are available upon reasonable request to the authors.

## Acknowledgments

NYDA® for the treatment of affected persons was kindly provided by Pohl-Boskamp GmbH & Co KG. We thank the 96 patients for participating in the study and the village tungiasis health workers for their work. The project social workers, Mike Banalyaki and Gertrude Nabbale, together with the project nurse, Proscovia Mukobe, oversaw data collection in the field. We also thank Dr. Susanne Wiese for her work during earlier phases of this study. The project activities in Uganda were coordinated by Innovations for Tropical Diseases Elimination (IFOTRODE).

